# Real-World Clinicopathological Characteristics, Metastatic Distribution, and Treatment Patterns of Triple-Negative Breast Cancer: A Single-Institution Experience from Nigeria

**DOI:** 10.64898/2026.06.10.26355358

**Authors:** Anthonia Chima Sowunmi, Chidi Agbakwuru, Eben Aje, Kehinde Ololade, Temitope Andero, Chukwuebuka Godspower Eze, Bukola Oshikanlu

## Abstract

**Background:** Triple-negative breast cancer (TNBC) is an aggressive breast cancer subtype associated with poor prognosis and a disproportionate disease burden among women of African ancestry. Contemporary data describing its clinicopathologic characteristics, metastatic patterns, treatment, and factors associated with advanced presentation in sub-Saharan Africa remain limited.

**Methods:** We conducted a retrospective cohort study of 869 women with histologically confirmed TNBC managed at a major tertiary cancer centre in Nigeria between May 2019 and December 2025. Demographic, clinicopathologic, treatment, and metastatic data were extracted from institutional electronic medical records. Advanced-stage presentation was defined as American Joint Committee on Cancer (AJCC) Stage III–IV disease, and multivariable logistic regression was performed to identify factors independently associated with advanced-stage presentation.

**Results:** The mean age at diagnosis was 52.1 ± 12.3 years. Most patients presented with Stage III (42.9%) or Stage IV (35.1%) disease, while only 21.9% presented with Stage I–II disease. Grade II (51.3%) and Grade III (36.5%) tumours predominated, and invasive ductal carcinoma accounted for 83.0% of cases. Among 305 patients with Stage IV disease, lung, bone, liver, and brain metastases occurred in 48.5%, 38.7%, 20.0%, and 18.4%, respectively. Overall, 73.1% underwent surgery, 83.2% received chemotherapy, and 63.5% received radiotherapy. On multivariable analysis, tumour grade was independently associated with advanced-stage presentation. Compared with Grade I tumours, Grade II tumours were associated with significantly greater odds of advanced-stage disease (adjusted odds ratio [aOR] 2.45, 95% confidence interval [CI] 1.39–4.32; p = 0.002), while Grade III tumours demonstrated more than five-fold higher odds (aOR 5.15, 95% CI 2.73–9.82; p < 0.001).

**Conclusion:** These findings demonstrate that TNBC in this Nigerian cohort presented predominantly at advanced stages and showed a visceral, lung-predominant metastatic distribution. The independent association between higher tumour grade and advanced-stage presentation underscores the need for earlier diagnosis, appropriate staging pathways, and strengthened multidisciplinary management in resource-constrained settings.

## Introduction

Breast cancer remains the most frequently diagnosed malignancy among women globally and a leading cause of cancer-related mortality, with an estimated 2.3 million new cases and over 666,000 deaths recorded worldwide in 2022.^1^ The burden is not evenly distributed. Africa carries a disproportionate share of breast cancer mortality relative to its incidence, a pattern captured by the region’s markedly elevated mortality-to-incidence ratio compared with high-income settings.^1^ Within this landscape, Nigeria stands out starkly: it recorded the highest absolute number of breast cancer cases and deaths on the continent in 2022, alongside one of the highest age-standardised mortality rates in Africa.^2^ Projections extending to 2050 suggest that this burden will only intensify, with sub-Saharan Africa expected to experience some of the steepest rises in both incidence and mortality worldwide, driven by population growth, ageing, and persistent gaps in early detection infrastructure.^3^

Triple-negative breast cancer (TNBC), defined immunohistochemically by the absence of estrogen receptor, progesterone receptor, and HER2 overexpression, occupies a particularly troubling position within this context. It is clinically and biologically the most aggressive of the major breast cancer subtypes, characterised by rapid proliferation, early distant recurrence, and a truncated interval between relapse and death compared with hormone receptor-positive disease.^4^ Despite major advances in immunotherapy, PARP inhibition and antibody-drug conjugates, access to these therapies remains highly unequal globally, and cytotoxic chemotherapy continues to underpin treatment in many resource-constrained settings.^5^ In Nigerian populations, TNBC has also been characterised by adverse pathological features. Tissue microarray-based evaluation of Nigerian breast cancers demonstrated a predominance of high-grade triple-negative phenotypes, reinforcing concerns regarding the biologically aggressive clinicopathologic profile of TNBC in this population.^6^

This heterogeneity takes on particular significance in populations of African ancestry, where TNBC occurs at a substantially higher frequency than in women of European descent. Institutional series from Nigeria have repeatedly documented this predilection: tissue microarray-based taxonomy of Nigerian breast cancers has shown a predominance of high-grade triple-negative phenotypes,^6^ while subsequent immunohistochemical surveys from Lagos^7^ and clinicopathologic reviews from Kano^8^ have reinforced the impression of a disease that is not only more common but also biologically more adverse in this population, frequently presenting in younger, pre-menopausal women. Whether this reflects genuine differences in tumour biology, differential access to screening and diagnosis, or an interplay of both remains an area of active investigation.

Compounding the biological aggressiveness of TNBC in this setting is the pattern of late-stage presentation that continues to characterise breast cancer care across sub-Saharan Africa. A systematic review and meta-analysis of stage at diagnosis across the region found that the majority of women present with locally advanced or metastatic disease, in sharp contrast to the early-stage predominance seen in high-income countries.^9^ Structural barriers, including limited screening infrastructure, delays in specialist referral, and constrained access, have been repeatedly implicated in this pattern across African cancer care systems.^10^ Even within Nigeria’s better-resourced institutional databases, treatment delivery remains inconsistent, with substantial proportions of patients not completing guideline-concordant multimodal therapy.^11^

The clinical consequences of TNBC’s biological aggressiveness are perhaps most visible in its pattern of distant spread. TNBC exhibits a distinct organotropism relative to other breast cancer subtypes, with a disproportionate predilection for visceral organs, the central nervous system, and bone, rather than the more indolent bone-predominant pattern typical of luminal disease.^12^ This metastatic behaviour, combined with the scarcity of targeted agents, continues to shape prognosis: even as global therapeutic advances, including immune checkpoint blockade and antibody-drug conjugates, have begun to alter outcomes in early and metastatic TNBC in well-resourced settings, the translation of these advances into routine practice across sub-Saharan Africa remains limited.^13^

Despite the disproportionate burden of TNBC in Nigerian women, comprehensive, contemporary institutional data describing its full clinicopathologic spectrum, patterns of metastatic spread, and treatment delivery within a single high-volume Nigerian oncology centre remain scarce. Much of the existing literature is drawn from smaller pathology-based series or focuses narrowly on receptor expression profiling, rather than integrating staging, metastatic patterns, and treatment patterns within one cohort. This study was therefore designed to describe the clinicopathologic characteristics, metastatic patterns, and treatment patterns of TNBC among patients managed at a high-volume tertiary oncology referral centre in Lagos, Nigeria, and to identify factors associated with advanced-stage disease.

## Materials and Methods

### Study Design and Setting

This retrospective cohort study was conducted at a tertiary cancer center in Nigeria. The centre is a high-volume tertiary oncology referral facility providing multidisciplinary cancer care, including medical, surgical, and radiation oncology, pathology, diagnostic imaging, and palliative care services. It serves as a major referral centre for Lagos State and neighbouring states across southwestern Nigeria. The study evaluated the clinicopathologic characteristics, metastatic patterns, treatment modalities, and factors associated with advanced-stage presentation among patients with triple-negative breast cancer (TNBC) managed in routine clinical practice.

### Study Population and Eligibility Criteria

The study population comprised patients with histologically confirmed invasive breast carcinoma managed at the centre between May 2019 and December 2025. Eligible patients had triple-negative breast cancer, defined by the absence of oestrogen receptor (ER), progesterone receptor (PR), and human epidermal growth factor receptor 2 (HER2) expression based on institutional immunohistochemistry (IHC) reports. ER and PR negativity were defined as nuclear staining in <1% of tumour cells, while HER2 negativity was determined according to the final institutional pathology interpretation, comprising IHC scores of 0 or 1+, or non-amplified ISH/FISH results for IHC 2+ cases. Patients with confirmed TNBC were included irrespective of disease stage at presentation. Patients with incomplete receptor characterisation, duplicate records, recurrent disease without baseline clinicopathologic information, or records lacking essential variables required for the primary analyses were excluded. The final study cohort consisted of 869 patients.

### Data Sources and Data Collection

Clinical data were retrospectively extracted from the institutional electronic medical record system using a standardised data abstraction form. Information was obtained from demographic records, outpatient and inpatient clinical notes, pathology and immunohistochemistry reports, radiological staging reports, multidisciplinary team documentation, operative records, chemotherapy treatment records, and radiotherapy planning systems.

Data abstraction was performed by trained research personnel under the supervision of a senior clinical investigator. Extracted data were cross-checked against multiple source documents where available to improve completeness and accuracy before being de-identified and stored in a secure password-protected research database.

### Data Management and Variable Definitions

Prior to analysis, the dataset underwent comprehensive cleaning and harmonisation using R statistical software (version 4.6.1; R Foundation for Statistical Computing, Vienna, Austria). Free-text entries, abbreviations, and synonymous clinical terminology were standardised using predefined coding dictionaries.

Disease stage was harmonised according to the American Joint Committee on Cancer (AJCC) 8th edition anatomic staging system and categorised as Stage I, II, III, or IV. Anatomic staging was used because prognostic stage incorporates tumour grade, which was evaluated separately in the regression model.

Tumour grade was classified according to the Nottingham histologic grading system as Grade I, II, or III. Histologic subtype was categorised as invasive ductal carcinoma, invasive lobular carcinoma, or other histologies. Treatment variables included surgery, chemotherapy, and radiotherapy. Chemotherapy regimens were categorised as anthracycline-based, platinum-containing, anthracycline-taxane, taxane-based, capecitabine-based, or other regimens. Radiotherapy variables included treatment site, fractionation schedule, and treatment technique (three-dimensional conformal radiotherapy [3D-CRT], intensity-modulated radiotherapy [IMRT], or volumetric modulated arc therapy [VMAT]).

### Outcome Definitions

The primary outcome was advanced-stage disease at initial presentation, defined as AJCC Stage III or Stage IV disease. The secondary outcome was the distribution of distant metastatic sites among patients presenting with Stage IV disease. Metastatic sites were identified from radiological and pathological reports and included lung, bone, liver, and brain metastases. Multiple-site metastasis was defined as documented involvement of two or more distant anatomical sites.

### Statistical Analysis

All statistical analyses were performed using R statistical software version 4.6.1 (R Foundation for Statistical Computing, Vienna, Austria). Prior to analysis, the dataset underwent comprehensive cleaning, validation, and harmonisation. Missing values in variables used for descriptive analyses were addressed using multiple imputation by chained equations (MICE). Twenty imputed datasets were generated using the mice package in R, and descriptive estimates were derived across the imputed datasets. Continuous variables were summarised using means with standard deviations or medians with ranges, as appropriate, while categorical variables were summarised using frequencies and percentages.

To identify factors independently associated with advanced-stage presentation, multivariable binary logistic regression was performed with advanced-stage disease (Stage III–IV) as the dependent variable. The regression analysis used a complete-case approach, whereby only patients with complete data for the outcome and all covariates included in the model were analysed. Independent variables entered simultaneously into the model were age (continuous), marital status, family history of breast cancer, and tumour grade. Marital status was dichotomised into Married (reference category) and Not married, with the latter comprising patients who were single, divorced, separated, or widowed.

Adjusted odds ratios (aORs) with corresponding 95% confidence intervals (CIs) were reported. Multicollinearity was assessed using variance inflation factors (VIFs). Statistical significance was defined as a two-sided p-value <0.05.

## Results

### Sociodemographic Characteristics of Patients with Triple-Negative Breast Cancer

A total of 869 women with triple-negative breast cancer (TNBC) were included in the study. The mean age at diagnosis was 52.1 ± 12.3 years, with a median age of 51 years (range: 22–97 years). Patients aged 40–49 years constituted the largest age group (29.7%), closely followed by those aged 50–59 years (29.5%). Women younger than 40 years accounted for 14.7% of the cohort, while 17.3% were aged 60–69 years and 8.9% were aged 70 years or older. Most patients were married (79.1%), followed by widowed (9.3%), single (8.5%), divorced (2.1%), and separated (1.0%). Employment status showed that 42.1% of patients were self-employed, 41.3% were formally employed, 10.5% were retired, and 6.1% were unemployed.

With respect to ethnicity, Yoruba patients constituted the largest proportion of the cohort (55.2%), followed by Igbo (23.5%), other ethnic groups (19.0%), and Hausa (2.3%). The “Others” category comprised minority ethnic groups and patients whose ethnicity was not documented. Christianity was the predominant religion, accounting for 80.3% of the cohort, followed by Islam (12.0%) and other religions (7.7%).

Regarding state of residence, most patients resided in Lagos State (68.9%). Other frequently represented states included Ogun (7.9%), Edo (4.7%), Delta (2.9%), Ondo (2.5%), Oyo (2.5%), and Rivers (2.5%), while each of the remaining states contributed less than 2% of the study population (Table 1).

**Table 1.**
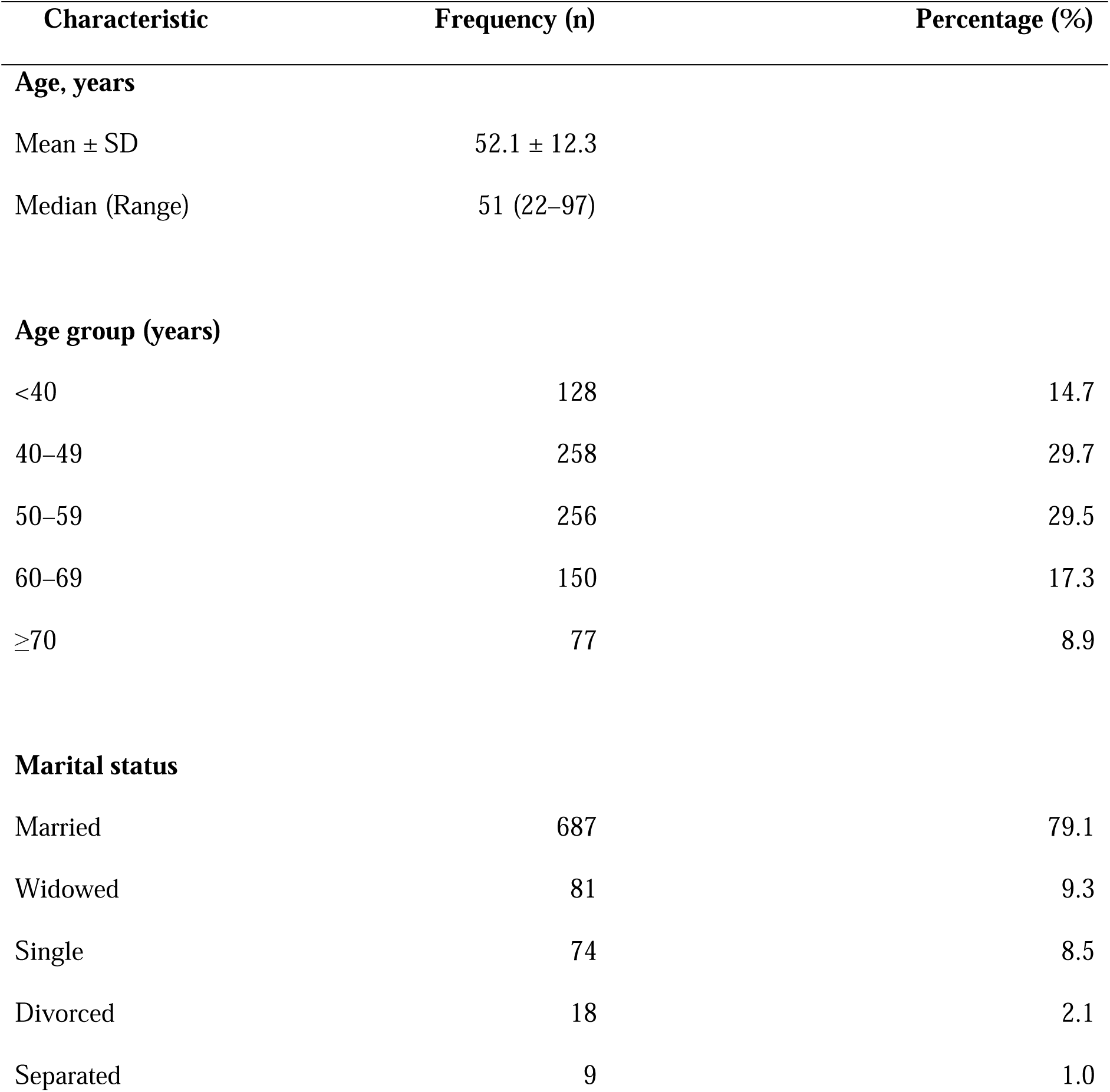
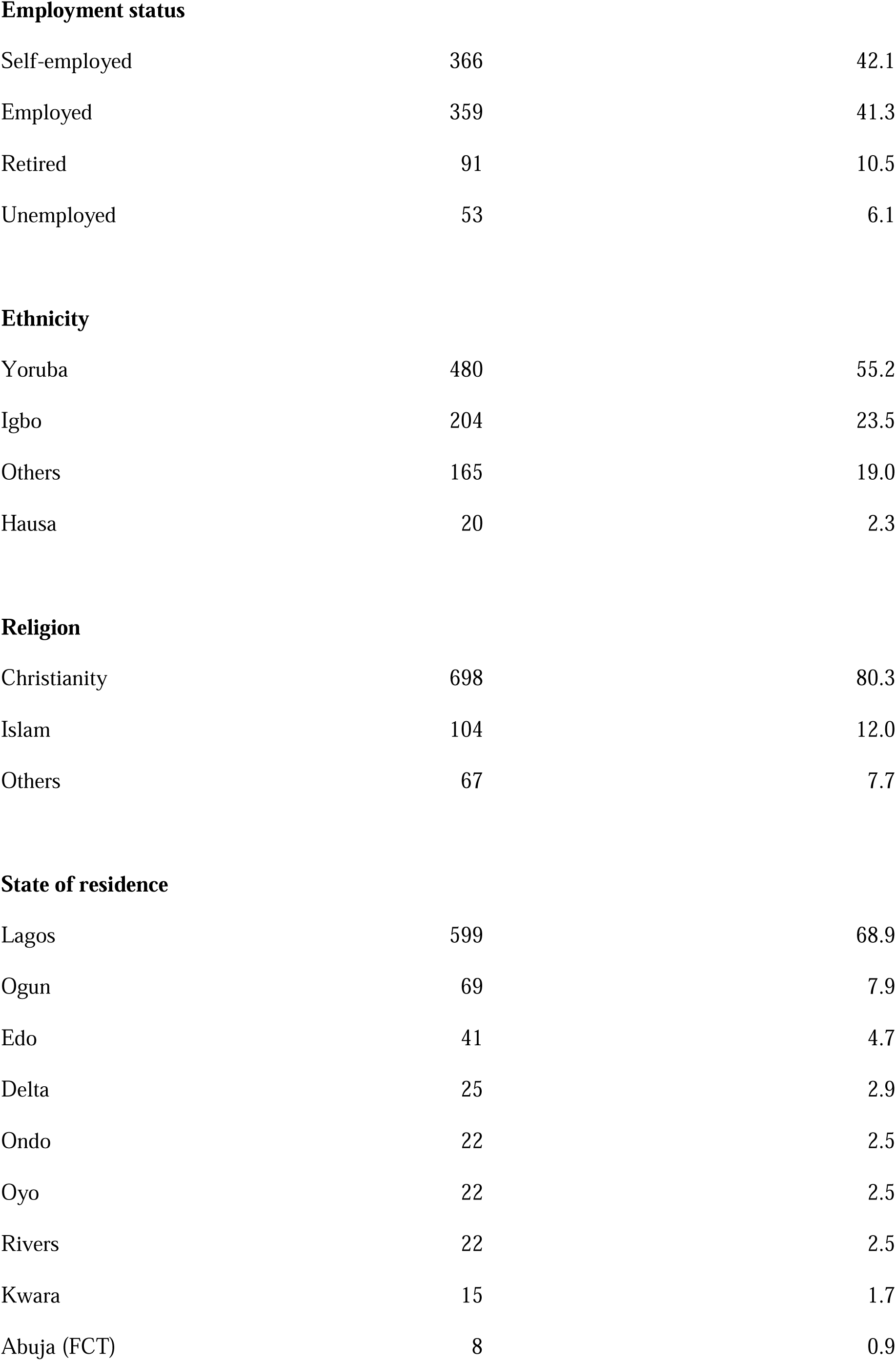
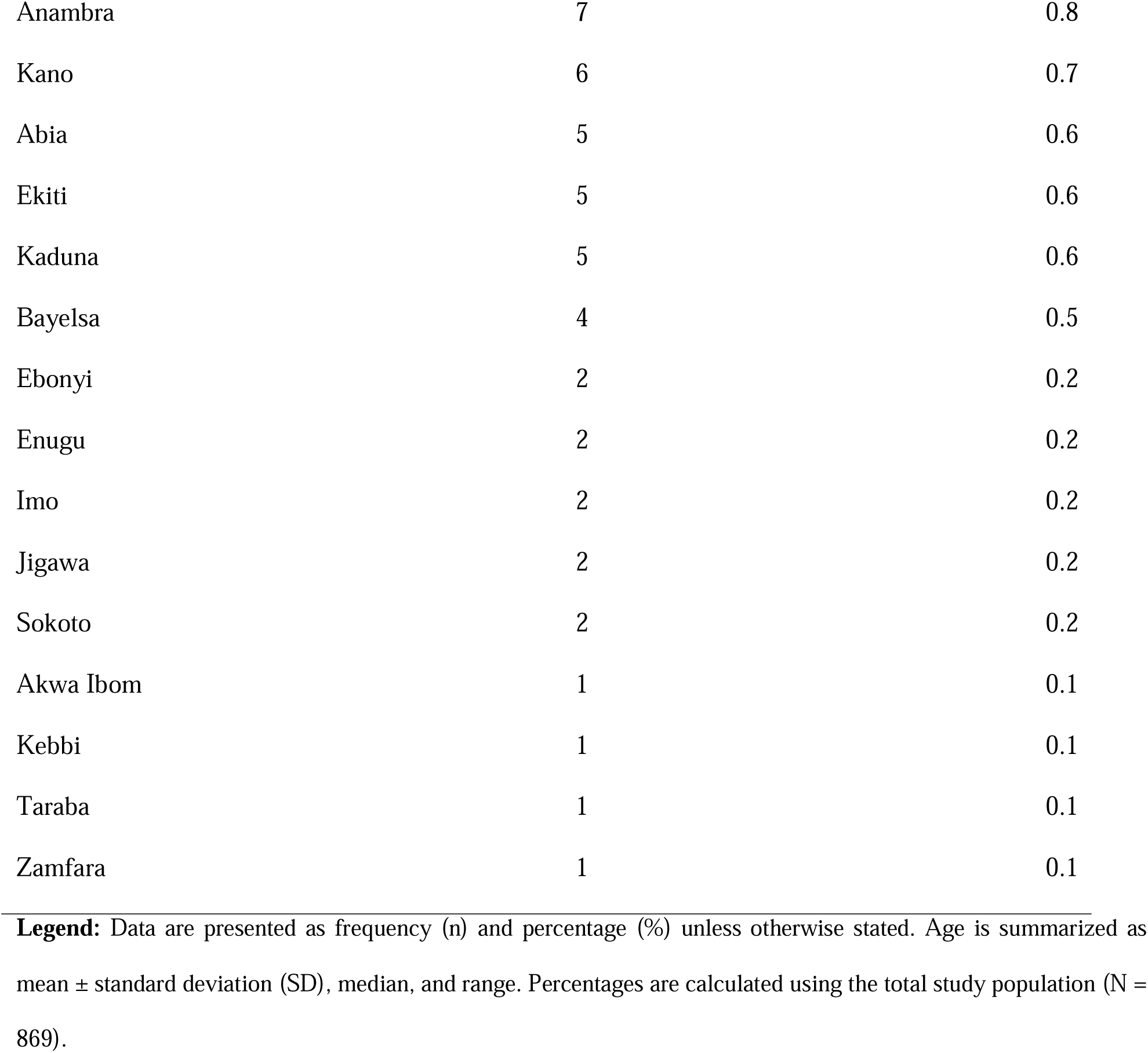
Sociodemographic characteristics of patients with triple-negative breast cancer (N = 869)

| Characteristic | Frequency (n) | Percentage (%) |
| --- | --- | --- |
| **Age, years** |  |  |
| Mean ± SD | 52.1 ± 12.3 | |
| Median (Range) | 51 (22–97) |  |
| **Age group (years)** |  |  |
| <40 | 128 | 14.7 |
| 40–49 | 258 | 29.7 |
| 50–59 | 256 | 29.5 |
| 60–69 | 150 | 17.3 |
| $\geq 70$ | 77 | 8.9 |
| **Marital status** |  |  |
| Married | 687 | 79.1 |
| Widowed | 81 | 9.3 |
| Single | 74 | 8.5 |
| Divorced | 18 | 2.1 |
| Separated | 9 | 1.0 |
| **Employment status** |  |  |
| Self-employed | 366 | 42.1 |
| Employed | 359 | 41.3 |
| Retired | 91 | 10.5 |
| Unemployed | 53 | 6.1 |
| **Ethnicity** |  |  |
| Yoruba | 480 | 55.2 |
| Igbo | 204 | 23.5 |
| Others | 165 | 19.0 |
| Hausa | 20 | 2.3 |
| **Religion** |  |  |
| Christianity | 698 | 80.3 |
| Islam | 104 | 12.0 |
| Others | 67 | 7.7 |
| **State of residence** |  |  |
| Lagos | 599 | 68.9 |
| Ogun | 69 | 7.9 |
| Edo | 41 | 4.7 |
| Delta | 25 | 2.9 |
| Ondo | 22 | 2.5 |
| Oyo | 22 | 2.5 |
| Rivers | 22 | 2.5 |
| Kwara | 15 | 1.7 |
| Abuja (FCT) | 8 | 0.9 |
| Anambra | 7 | 0.8 |
| Kano | 6 | 0.7 |
| Abia | 5 | 0.6 |
| Ekiti | 5 | 0.6 |
| Kaduna | 5 | 0.6 |
| Bayelsa | 4 | 0.5 |
| Ebonyi | 2 | 0.2 |
| Enugu | 2 | 0.2 |
| Imo | 2 | 0.2 |
| Jigawa | 2 | 0.2 |
| Sokoto | 2 | 0.2 |
| Akwa Ibom | 1 | 0.1 |
| Kebbi | 1 | 0.1 |
| Taraba | 1 | 0.1 |
| Zamfara | 1 | 0.1 |

**Legend:** Data are presented as frequency (n) and percentage (%) unless otherwise stated. Age is summarized as mean ± standard deviation (SD), median, and range. Percentages are calculated using the total study population (N = 869).

### Clinicopathologic Characteristics

The clinicopathologic characteristics of the 869 patients with triple-negative breast cancer are presented in Table 2. Stage III disease was the most common stage at presentation, accounting for 373 patients (42.9%), followed by Stage IV disease in 305 patients (35.1%) and Stage II disease in 149 patients (17.1%). Early-stage disease was relatively uncommon, with 42 patients (4.8%) presenting with Stage I disease.

**Table 2.** Clinicopathologic Characteristics of Patients with Triple-Negative Breast Cancer.

| Variable | Category | n | % |
| --- | --- | --- | --- |
| **Stage** | Stage I | 42 | 4.8 |
|  | Stage II | 149 | 17.1 |
|  | Stage III | 373 | 42.9 |
|  | Stage IV | 305 | 35.1 |
| **Tumour grade** | Grade I | 106 | 12.2 |
|  | Grade II | 446 | 51.3 |
|  | Grade III | 317 | 36.5 |
| **Histology** | Invasive ductal carcinoma | 721 | 83.0 |
|  | Invasive lobular carcinoma | 23 | 2.6 |
|  | Other histologies | 125 | 14.4 |
| **Menopausal status** | Pre-menopausal | 250 | 28.8 |
|  | Peri-menopausal | 60 | 6.9 |
|  | Post-menopausal | 559 | 64.3 |
| **Family history of breast cancer** | No | 728 | 83.8 |
|  | Yes | 141 | 16.2 |
| **Laterality** | Bilateral | 23 | 2.6 |
|  | Left | 424 | 48.8 |
|  | Right | 422 | 48.6 |

**Legend:** Clinicopathologic characteristics of patients with triple-negative breast cancer. Values are presented as frequency (n) and percentage (%). Percentages were calculated using the total study population (N = 869). Histologic subtypes with low frequencies were grouped under "Other histologies." Percentages may not sum exactly to 100% because of rounding.

Tumour grade was predominantly Grade II, occurring in 446 patients (51.3%), while 317 patients (36.5%) had Grade III tumours and 106 patients (12.2%) had Grade I tumours. Invasive ductal carcinoma was the predominant histologic subtype, accounting for 721 patients (83.0%), followed by other histologic subtypes in 125 patients (14.4%) and invasive lobular carcinoma in 23 patients (2.6%).

Post-menopausal women constituted the largest proportion of the cohort (559 patients; 64.3%), followed by pre-menopausal (250 patients; 28.8%) and peri-menopausal women (60 patients; 6.9%). Most patients (728; 83.8%) had no documented family history of breast cancer, whereas 141 (16.2%) reported a positive family history.

Tumour laterality was nearly equally distributed between the left and right breasts. Left-sided tumours were observed in 424 patients (48.8%), while 422 patients (48.6%) had right-sided disease. Bilateral breast involvement was uncommon, occurring in 23 patients (2.6%).

### Distribution of Metastatic Sites Among Patients with Stage IV Disease

The distribution of metastatic sites among patients with Stage IV triple-negative breast cancer is presented in Figure 1. Lung was the most common metastatic site, occurring in 148 patients (48.5%), bone metastasis was documented in 118 patients (38.7%), while liver and brain metastases occurred in 61 (20.0%) and 56 (18.4%) patients, respectively.

**Figure 1.**
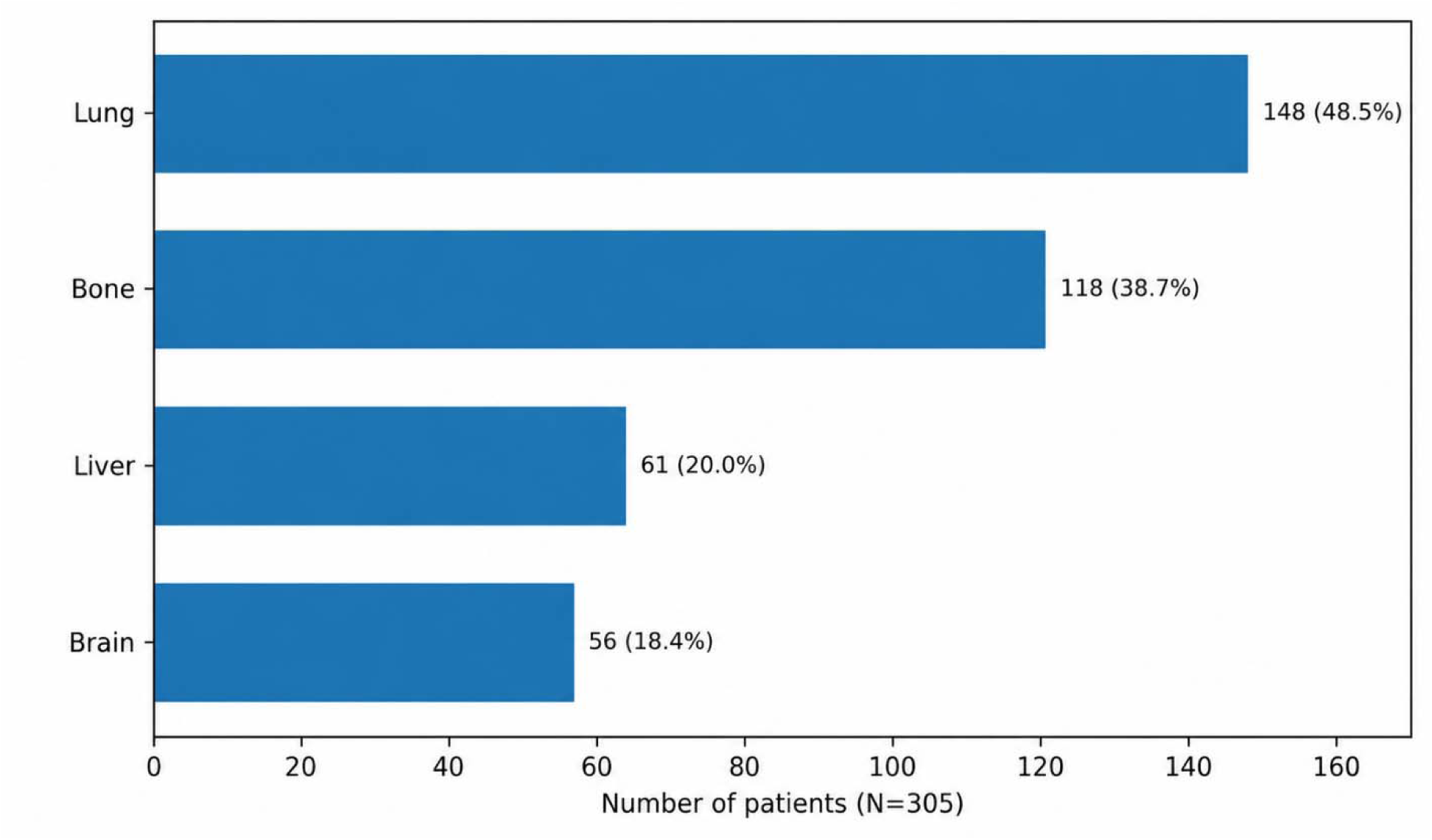
Distribution of Metastatic Sites Among Patients with Stage IV Triple-Negative Breast Cancer (N = 305) **Legend:** Horizontal bar chart illustrating the distribution of documented distant metastatic sites among patients with Stage IV triple-negative breast cancer (N = 305). Values are presented as frequency (n) and percentage (%). Percentages were calculated using the total number of patients with Stage IV disease as the denominator. Because patients could have metastases involving more than one distant anatomical site, percentages do not sum to 100%.

### Treatment Patterns

Overall, 635 patients (73.1%) underwent surgical treatment, of whom 450 (70.9%) received mastectomy and 185 (29.1%) underwent breast-conserving surgery. Chemotherapy was administered to 723 patients (83.2%), with anthracycline-based regimens being the most frequently prescribed (41.8%), followed by platinum-containing regimens (28.8%) and anthracycline–taxane combinations (16.3%). Radiotherapy was delivered to 552 patients (63.5%). Among those who received radiotherapy, the chest wall was the most commonly irradiated site, accounting for 60.0% of treatment courses, followed by the spine (15.2%), breast (14.7%), other sites (12.9%), and brain (12.3%). Intensity-modulated radiotherapy (IMRT, 48.6%) was the predominant treatment technique, followed by 3D-conformal radiotherapy (31.2%) and volumetric modulated arc therapy (20.3%). The fractionation schedule most frequently employed was moderately hypofractionated breast radiotherapy (42–43 Gy in 15–16 fractions), used in 54.3% of radiotherapy courses, while hypofractionated regimens of 20 Gy in 5 fractions accounted for 23.6%. Detailed treatment patterns are presented in Table 3.

**Table 3.** Treatment Patterns Among Patients with Triple-Negative Breast Cancer.

| Domain | Category | N | % |
| --- | --- | --- | --- |
| **Surgery (n = 869)** | Yes | 635 | 73.1 |
|  | No | 234 | 26.9 |
| **Type of surgery (n = 635)** | Mastectomy | 450 | 70.9 |
|  | Lumpectomy/BCS | 185 | 29.1 |
| **Chemotherapy (n = 869)** | Yes | 723 | 83.2 |
|  | No | 146 | 16.8 |
| **Chemotherapy regimen (n = 723)** | Anthracycline-based | 302 | 41.8 |
|  | Platinum-containing | 208 | 28.8 |
|  | Anthracycline–taxane | 118 | 16.3 |
|  | Taxane-based | 54 | 7.5 |
|  | Capecitabine-based | 41 | 5.7 |
| **Radiotherapy (n = 869)** | Yes | 552 | 63.5 |
|  | No | 317 | 36.5 |
| **RT site distribution among patients receiving radiotherapy (n = 552)** | Chest wall | 331 | 60.0 |
|  | Spine | 84 | 15.2 |
|  | Breast | 81 | 14.7 |
|  | Other sites | 71 | 12.9 |
|  | Brain | 68 | 12.3 |
| **RT fractionation schedules among patients receiving radiotherapy (n = 552)** | 42–43 Gy / 15–16 fractions | 300 | 54.3 |
|  | 20 Gy / 5 fractions | 130 | 23.6 |
|  | 30 Gy / 10 fractions | 56 | 10.1 |
|  | 45 Gy / 18 fractions | 22 | 4.0 |
|  | 40 Gy / 15 fractions | 14 | 2.5 |
|  | Others | 113 | 20.5 |
| **RT technique (n = 552)** | IMRT | 268 | 48.6 |
|  | 3D-CRT | 172 | 31.2 |
|  | VMAT | 112 | 20.3 |

**Legend:** Values are presented as frequency (n) and percentage (%). Percentages for surgery, chemotherapy, and radiotherapy (yes/no) were calculated using the total study population (N = 869). Percentages for surgery type, chemotherapy regimen, and radiotherapy technique were calculated using the corresponding treatment-specific denominators (n = 635, 723, and 552, respectively). For radiotherapy site distribution and fractionation, percentages were calculated using the total number of radiotherapy treatment courses (n = 552) as the denominator. These categories are not mutually exclusive because individual patients may have undergone irradiation to more than one anatomical site and/or received more than one radiotherapy fractionation schedule during the course of their treatment. Consequently, the sum of percentages for these categories exceeds 100%.

### Factors Associated with Advanced-Stage Presentation: Multivariable Logistic Regression

A multivariable logistic regression model was fitted to identify factors independently associated with advanced-stage (Stage III–IV) presentation among patients with triple-negative breast cancer. The model adjusted for age, marital status (married versus not married), family history of breast cancer, and tumour grade. Adjusted odds ratios (aORs), 95% confidence intervals (CIs), and p-values are presented in Table 4. Increasing tumour grade remained independently associated with advanced-stage presentation. Compared with patients with Grade I tumours, those with Grade II disease had approximately 2.5-fold higher odds of presenting with advanced-stage disease (aOR = 2.45, 95% CI: 1.39–4.32; *p* = 0.002), while Grade III tumours were associated with more than a five-fold increase in the odds of advanced-stage presentation (aOR = 5.15, 95% CI: 2.73–9.82; *p* < 0.001). Age, marital status, and family history of breast cancer were not independently associated with advanced-stage presentation after adjustment.

**Table 4.** Multivariable logistic regression analysis of factors associated with advanced-stage triple-negative breast cancer.

| Variable | Category | Adjusted OR | 95% CI | *p*-value |
| --- | --- | --- | --- | --- |
| Age (years) | Per one-year increase | 1.01 | 1.00–1.03 | 0.116 |
| Marital Status | Married | Reference | — | — |
|  | Not married | 1.22 | 0.72–2.17 | 0.476 |
| Family History of Breast Cancer | No | Reference | — | — |
|  | Yes | 1.23 | 0.69–2.26 | 0.496 |
| Tumour Grade | Grade I | Reference | — | — |
|  | Grade II | 2.45 | 1.39–4.32 | 0.002 |
|  | Grade III | 5.15 | 2.73–9.82 | <0.001 |

**Legend:** Multivariable logistic regression analysis of factors independently associated with advanced-stage (Stage III–IV) presentation among patients with triple-negative breast cancer. The model included age, marital status, family history of breast cancer, and tumour grade using complete-case analysis. Marital status was dichotomised into **Married** (reference category) and **Not married**, with the latter comprising participants who were single, divorced, separated, or widowed. Adjusted odds ratios (aORs) greater than 1 indicate increased odds of presenting with advanced-stage disease relative to the reference category. Statistical significance was defined as $p < 0.05$ .

## Discussion

This retrospective cohort of 869 women with triple-negative breast cancer (TNBC) managed at the Cancer Centre represents one of the largest single-institution, TNBC-specific series reported from Nigeria to date. The integrated view of sociodemographic profile, clinicopathologic characteristics, metastatic behaviour, treatment delivery, and factors associated with advanced-stage presentation within a single clinical population offers a comprehensive snapshot of the disease burden in a high-volume tertiary referral centre in sub-Saharan Africa. The findings largely reaffirm the aggressive phenotype and late-presenting pattern of TNBC previously described in Nigerian and broader African settings, while also surfacing a few patterns that diverge from expectation and merit closer interrogation.

The mean age at diagnosis in this cohort was 52.1 ± 12.3 years, with women aged 40 to 49 years and 50 to 59 years together accounting for nearly 60% of the population (Table 1). This is somewhat older than the mean age of 49.3 years reported in the Lagos University Teaching Hospital TNBC series^7^ and serves as an important reminder that although TNBC in women of African ancestry occurs at a younger age than in women of European descent, this predilection represents a shift in the population distribution rather than a phenomenon confined exclusively to the very young.^14^ The proportion of patients below 40 years (14.7%) in this series is lower than some smaller Nigerian hospital-based reports, such as the Kano clinicopathologic review where younger patients constituted a larger fraction.^8^ This discrepancy likely reflects differences in referral patterns or catchment population rather than a genuine departure from regional epidemiology, given that the Centre draws from a broad, multi-state referral base across southwestern Nigeria. The predominance of married patients (79.1%) and the combined proportion of self-employed or formally employed women (83.4%) mirror the socioeconomic profile described elsewhere in Nigerian oncology populations and carry implications for the affordability and completion of sustained multimodal treatment.^15^ The stage distribution in this cohort, with Stage III (42.9%) and Stage IV (35.1%) disease together accounting for nearly four out of every five patients at presentation (Table 2), sits at the advanced end of what has been documented nationally. A recent meta-analysis of Nigerian stage-at-diagnosis data spanning 2018 to 2023 found only a modest shift in stage distribution compared with the 2000 to 2018 period, with a slight increase rather than decrease in Stage III presentation, suggesting that national interventions aimed at promoting earlier presentation have so far achieved limited measurable impact.^15^ Because the present cohort is restricted to TNBC, a receptor subtype independently associated with more advanced stage at presentation in several African series,^6,8^ the gap between this cohort’s stage distribution and all-comer national estimates is not entirely unexpected. However, it underscores how disproportionately this receptor subtype concentrates the burden of late-stage disease.

The grade distribution, dominated by Grade II (51.3%) and Grade III (36.5%) tumours, is consistent with the well-established observation that TNBC is enriched for high-grade, high-proliferation histology relative to hormone receptor-positive disease^5^ and aligns with earlier Nigerian pathology-based series that similarly documented a predominance of high-grade phenotypes among triple-negative tumours.^6^ Histological grade is more than a descriptive label; it is a recognised surrogate for the underlying proliferative activity and genomic instability that drive clinical aggressiveness,^16^ a point that becomes directly relevant when the regression findings are considered. Two findings in the clinicopathologic profile depart somewhat from prior expectation and warrant discussion. The menopausal distribution, with post-menopausal women comprising 64.3% of the cohort (Table 2), sits at odds with the frequently cited observation that TNBC clusters preferentially among pre-menopausal women of African ancestry.^6,7^ This likely reflects, at least in part, the overall older age structure of this cohort relative to smaller Nigerian series, and serves as a useful corrective to the notion that pre-menopausal clustering is so dominant as to make post-menopausal TNBC an unusual finding in a large, relatively unselected clinical population. The low documented rate of positive family history (16.2%) should also be interpreted cautiously. While broadly consistent with figures from other Nigerian cohorts, it sits against a backdrop in which genetic counselling and BRCA1/2 testing remain minimally integrated into routine Nigerian oncology practice. A 2025 survey of Nigerian healthcare providers found that only about a third had ever requested genetic testing for a patient, with virtually all such testing conducted through private, out-of-pocket arrangements rather than routine institutional pathways.^17^ Given that pathogenic BRCA1/2 variants are increasingly recognised, though still under-characterised, in Nigerian and other sub-Saharan African populations,^18^ it is plausible that the true prevalence of hereditary predisposition in this cohort is under-ascertained rather than genuinely low, a distinction with direct implications for how the family history variable in the regression model should be interpreted.

The burden of advanced-stage disease in this cohort was substantial, with Stage III and Stage IV disease accounting for 42.9% and 35.1% of patients, respectively (Table 2). Among the 305 patients who presented with Stage IV disease, the lung was the most frequent site of distant metastasis (48.5%), followed by bone (38.7%) (Figure 1). Liver and brain metastases occurred in 20.0% and 18.4% of patients, respectively. This metastatic distribution is broadly consistent with the recognised visceral-predominant organotropism of TNBC, which contrasts with the bone-predominant pattern typical of luminal, hormone receptor-positive disease.^12^ Large population-based analyses have consistently demonstrated that molecular subtype influences metastatic organotropism, with hormone receptor-positive disease showing a predilection for bone metastases, whereas TNBC more frequently metastasises to visceral organs, particularly the lung.^19^ TNBC-specific analyses from the Surveillance, Epidemiology, and End Results (SEER) programme similarly identify the lung as the predominant site of distant spread.^20,21^ Although a recent Ghanaian radiotherapy-centre cohort reported bone as the commonest metastatic site, narrowly exceeding lung involvement,^22^ that study included all breast cancer molecular subtypes rather than TNBC alone, likely accounting for the greater prominence of skeletal metastases. The concordance between the present TNBC-specific findings and the wider international literature suggests that the biological determinants of metastatic organotropism in TNBC remain largely conserved across populations despite marked differences in healthcare systems and stage at presentation.

Surgical treatment was delivered to 73.1% of patients, with mastectomy accounting for more than two-thirds of surgical procedures performed (70.9%) against only 29.1% breast-conserving surgery (Table 3). This mastectomy-dominant pattern is consistent with the wider Nigerian surgical literature; a ten-year review of breast cancer surgery at University College Hospital, Ibadan, similarly found surgical decision-making shaped primarily by advanced tumour stage rather than by surgical preference or technical capacity.^23^ A multi-country sub-Saharan African study of breast cancer care found mastectomy rates of 64% to 67% against breast-conserving rates of only 15% to 26% across Ghana, Kenya, and Nigeria.^24^ The relatively low rate of breast-conserving surgery in this cohort reflects the advanced stage at presentation, where breast conservation is often not feasible. Expanding breast-conserving surgery services should be a priority for quality improvement, particularly for patients with early-stage disease who could benefit from less extensive surgery. Chemotherapy was administered to the large majority of patients (83.2%), with anthracycline-based regimens most frequently used (41.8%), followed by platinum-containing regimens (28.8%) and anthracycline-taxane combinations (16.3%). Radiotherapy was delivered to 63.5% of patients, with intensity-modulated radiotherapy (IMRT, 48.6%) being the predominant technique among those treated (Table 3). This is a comparatively favourable figure set against the same multi-country study, in which only 28% of Nigerian patients in the sampled cohort received radiotherapy at all,^24^ and lends support to this centre’s description as a well-equipped facility relative to the broader national infrastructure. The fractionation schedules show considerable heterogeneity, with the majority receiving moderately hypofractionated breast radiotherapy (42 to 43 Gy in 15 to 16 fractions), while 23.6% received hypofractionated regimens (20 Gy in 5 fractions), likely reflecting palliative treatment in the metastatic setting. The integration of IMRT and VMAT techniques indicates access to modern radiotherapy technology, though the clinical impact on toxicity and outcomes requires prospective evaluation.

The multivariable logistic regression findings (Table 4) reinforce the biological plausibility of the clinicopathologic patterns already described. Increasing tumour grade remained the only independent factor associated with advanced-stage disease after adjustment for age, marital status, and family history of breast cancer. Compared with Grade I tumours, Grade II tumours were associated with more than twice the odds of presenting with advanced-stage disease (aOR 2.45, 95% CI: 1.39 to 4.32), while Grade III tumours were associated with more than a five-fold increase in the odds of advanced-stage presentation (aOR 5.15, 95% CI: 2.73 to 9.82). These findings are consistent with the understanding of histological grade as a surrogate for proliferative activity and genomic instability, both of which favour more rapid disease progression and, plausibly, a shorter interval between tumour onset and clinical detection at an already advanced stage.^16^ The observation that tumour grade remained independently associated with advanced-stage presentation, despite adjustment for demographic and familial factors, further underscores its importance as a marker of disease aggressiveness within this TNBC cohort. Clinically, this finding supports a lower threshold for comprehensive baseline staging in patients with high-grade tumours, including dedicated chest imaging given the lung-predominant metastatic pattern observed in this study. Age was not independently associated with advanced-stage presentation (aOR 1.01 per year, p = 0.116), while neither marital status nor family history of breast cancer demonstrated statistically significant associations after adjustment. The absence of an independent association with family history should be interpreted cautiously. As noted earlier, the relatively low documented prevalence of positive family history and the limited availability of genetic counselling and BRCA1/2 testing in Nigeria likely contribute to substantial under-ascertainment of hereditary susceptibility, thereby attenuating any true association between familial predisposition and stage at presentation.^17,18^

## Limitations

Several limitations should be considered when interpreting these findings. First, the retrospective, single-centre design carries the usual limitations of this study type, including the likelihood of referral bias toward more advanced disease at a high-volume tertiary cancer centre, and constrained generalisability of these findings to primary or secondary-level facilities elsewhere in Nigeria. Second, survival ascertainment for this cohort is ongoing, and outcome-related calls to patients and next of kin had not been completed at the time of this analysis. This meant that survival analysis, which would have substantially strengthened the clinical and prognostic value of this work, could not be undertaken; this remains a priority for a planned follow-up analysis. Third, no molecular or genomic characterisation, including BRCA1/2 germline testing or TNBC intrinsic molecular subtyping, was available for this cohort, which limits the ability to relate the clinicopathologic and metastatic patterns observed here to the underlying tumour biology.^17,18^ Fourth, the absence of data on treatment completion, dose intensity, and treatment-related toxicities constrains the interpretation of treatment patterns and their impact on outcomes.

## Conclusion

Notwithstanding these limitations, this study offers a substantial and clinically grounded contribution to the understanding of triple-negative breast cancer in Nigeria. With 869 patients, it stands among the largest TNBC-specific cohorts reported from a single sub-Saharan African institution. The methodological approach, including thorough data cleaning, complete-case analysis, and multivariable regression modelling, strengthens confidence in the associations reported. The findings reaffirm that TNBC in this Nigerian population presents overwhelmingly at an advanced stage, that tumour grade is a robust independent factor associated with advanced presentation, and that its metastatic behaviour follows a visceral, lung-predominant pattern broadly consistent with the global TNBC organotropism literature despite the very different resource context in which this cohort was managed. These findings carry direct clinical implications, from reinforcing the case for early chest imaging in staging and surveillance protocols to highlighting the continued underutilisation of breast-conserving surgery and the near-complete absence of genetic counselling infrastructure as tractable targets for local quality improvement. As survival data from this cohort become available, they will allow these clinicopathologic and metastatic patterns to be anchored to prognosis, further consolidating the role of this centre in generating locally grounded, publication-quality evidence to guide TNBC care across Nigeria and globally.

## Data Availability

The minimal underlying dataset supporting the findings of this study is maintained securely by the corresponding author and can be made available upon reasonable request.

